# Longitudinal changes in optoretinography provide an early and sensitive biomarker of outer retinal disease

**DOI:** 10.1101/2025.03.12.25323718

**Authors:** Teng Liu, Benjamin Wendel, Jennifer Huey, Vimal Prabhu Pandiyan, Debarshi Mustafi, Jennifer R. Chao, Ramkumar Sabesan

## Abstract

**Objective:** To examine whether optoretinography (ORG) can provide greater sensitivity for assessing the time-course of disease progression in Retinitis Pigmentosa compared to standard clinical imaging in a longitudinal study,

**Design:** Cohort, longitudinal study.

**Participants:** Five non-syndromic RP patients and eight control subjects participated in the study.

**Methods:** Clinical examination, imaging sessions and data analysis were all conducted at the University of Washington. Five eyes of 5 patients diagnosed with RP, comparing standard clinical imaging to ORG, were collected over a 21-month span between August 2022 and May 2024.

**Main outcome and measures:** ORG response to visual stimuli, ellipsoid zone (EZ) width and outer segment length (OS length) were evaluated for longitudinal changes as markers of disease progression.

**Results:** The reduction in cone function with ORG over time exceeds that observed in standard clinical markers of photoreceptor structure - EZ width and OS length. EZ width and OSL decreased by 4.5% ± 5.9% and 6.5% ± 1.4%, respectively, approximately 9.9 and 6.9 times less than the reduction noted in ORG, respectively. The most notable degradation was noted at the borders of the transition zone, where ORG showed progressive and sub-clinical losses in photoreceptor function whereas standard OCT showed healthy, unaffected outer retinal structure.

**Conclusions:** Optoretinography detects sub-clinical disease and reliably identifies longitudinal markers of progression with greater sensitivity compared to standard clinical imaging. The ability to detect functional changes in the outer retina prior to standard clinical measures underscores its potential as a sensitive, accelerated and clinically-relevant outcome measure to guide patient selection and their therapeutic response in future clinical trials.

## Introduction

Sensitive biomarkers of retinal health are critical for monitoring disease progression and evaluating therapeutic response ^1,2^. Current clinical imaging tools are limited in their sensitivity and resolution to detect subtle degradation in retinal structure and function. This precludes early diagnosis and treatment, limiting visual recovery as there is a critical window for therapeutic intervention ^3^. Optical coherence tomography (OCT) and fundus autofluorescence (FAF) are hallmark structural measures of the outer retinal complex. Primary biomarkers include the ellipsoid band width on OCT ^4–8^ and hyper-autofluorescent (hyper-AF) rings on FAF ^9–12^. It remains unclear to what extent the remnant structure of photoreceptors still supports normal function. Furthermore, studies have shown that in advanced stages of disease, OCT and FAF reveal only minor changes over time, making it difficult to detect incremental progression or an accurate response to treatment ^13,14^. Clinical functional assessments, including BCVA ^15,16^, perimetry ^17^ and electroretinogram (ERG) ^18,19^, offer complementary information about retinal integrity. BCVA and perimetry represent the cumulative output of the entire visual system and are thus limited in their cellular specificity. Moreover, mechanisms in the retina and cortex that compensate for photoreceptor loss can allow patients to largely ignore visual symptoms until late in the disease process ^15,20,21^. The gold standard objective measure of retinal functional activity is ERG, but it lacks the spatial resolution to detect localized changes, particularly in advanced stages of disease ^22^. The urgent need for a non-invasive and sensitive biomarker of retinal disease progression has taken on renewed importance in the light of recent gene therapy clinical trials that have shown promise to restore outer retinal function ^23–28^.

Optoretinography (ORG) marks a significant advance in the non-invasive and sensitive monitoring of retinal structure and function *in vivo*. When implemented in an OCT, it provides a direct optical measure of light-evoked photoreceptor activity ^29–37^ in conjunction with the underlying three-dimensional structure responsible for the functional assay. The activity is linked to the initial steps in vision that originate in the outer retina - light absorption, photoisomerization, phototransduction and the visual cycle ^34,38^. A few recent studies have demonstrated the ability of ORG to detect sub- clinical changes in RP ^21,39,40^, particularly in the transition zone at the border of healthy and diseased areas, highlighting its potential for accurate disease assessment.

RP is the most common inherited retinal degeneration (IRD) and is the target of various therapeutic approaches ^23,41,42^. Whereas no approved treatments are available to halt or reverse the progression of RP, several clinical trials are underway ^43–45^. Clinical and high-resolution adaptive optics (AO) imaging have revealed the common outer retinal phenotypes in RP - reduction in EZ width ^6–8,11,46,47^, OS length ^48–50^ and cone density ^6,8^. Visual acuity ^51^ and perimetry ^17^ reveal progressive visual field constriction that eventually affects the central fovea. ERG ^19,52–54^ typically shows reduced or absent a- and b-waves, reflecting photoreceptor degeneration and inner retinal remodeling. Overall, these metrics show between 4-13 % annual rates of RP progression ^6,19,55^.

Here, in the first application of ORG to a longitudinal study, we test the hypothesis that cone dysfunction measured with ORG precedes the deterioration exhibited by standard clinical biomarkers in RP patients. We hypothesize that the ORG can detect photoreceptors in their earliest stages of outer segment compromise, at the border between the healthy-appearing central island and transition zone, where structural imaging suggests normal status. This retrospective longitudinal study spanned 12 to 21 months in 5 RP patients and compared ORG findings with EZ width and OS length in quantifying progression.

## Method

### Study participants

Five patients (ages 23–60 years-old (yo), mean: 33.4 yo) diagnosed with RP based on clinical presentation and relevant family history at the Karalis Johnson Retina Center, Department of Ophthalmology at University of Washington were enrolled in the study. All participants underwent genetic testing through CLIA-approved labs and four (RP02 – RP05) had a confirmed molecular genetic diagnosis (Table 1). Although genetic testing for RP01 was inconclusive, their clinical exam, imaging, Goldmann Visual Fields, and electrophysiology testing were consistent with RP. Research procedures followed the tenets of the Declaration of Helsinki. Eight age-matched normal controls were also enrolled in the study. Informed consent was obtained from all patients, and research protocols were approved by the institutional review board of the University of Washington (IRB#STUDY00002923). Patient characteristics are listed in Table 1.

**Table 1.** Characteristics of study participants.

| Patient ID | Age range | Sex | Genotype | Clinical presentation description | Follow-up duration (months) | Change of BCVA study eye (logMAR) | Reduction of EZ width (μm, %) |
| --- | --- | --- | --- | --- | --- | --- | --- |
| RP01 | 26-30 | F | Indeterminate | nyctalopia, myopia, "macular cysts" | 16, 20 | 0 | 310 (15%) |
| RP02 | 26-30 | M | *IMPDH1*<br>c.942_944del,<br>p.(Lys314del) | reduced peripheral vision, impaired dark adaptation | 14, 21 | 0.08 | 72 (5.3%) |
| RP03 | 56-60 | F | *RHO*<br>c.165C>A,<br>p.(Asn55Lys) | nyctalopia, increased glare while driving | 9, 15 | 0.10 | 130 (13.5%) |
| RP04 | 21-25 | F | *PRPF31*<br>c.323-1G>T | nyctalopia, high myopia, blurring of central vision with flashes | 6, 12 | -0.10 | 23 (0.9%) |
| RP05 | 26-30 | F | *RHO*<br>c.68C>A,<br>p.(Pro23His) | nyctalopia, reduced peripheral vision | 8, 16 | 0 | 34 (1.5%) |

Research imaging procedures were performed over three visits: baseline, first follow-up (f/u #1), and second follow-up (f/u #2), with a minimum of six months between each follow-up visit. The maximum length of follow-up ranged from 12 to 21 months. Cycloplegia was induced by 1% tropicamide ophthalmic solution for imaging.

The study involved standard clinical testing including OCT (Spectralis HRA+OCT system; Heidelberg Engineering, Vista, CA, USA), color fundus photography and fundus autofluorescence (FAF) (Optos, Marlborough, MA, USA). These conventional measures were compared to ORG (Figure 1).

**Figure 1.**
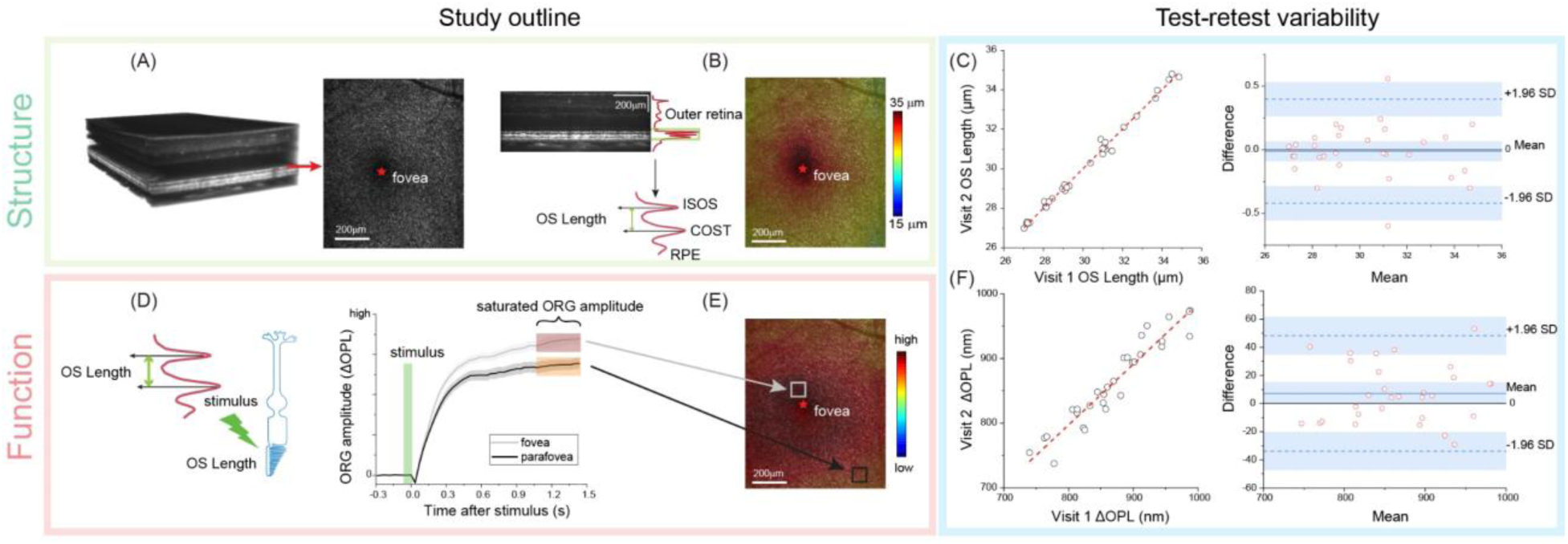
Overview of study design, illustrating the advanced structure-function assessment. OCT volumes along with the segmented *en face* image at the inner/outer segment-ellipsoid zone (ISOS-EZ) and outer segment tips (COST) help visualize the retinal structure (A). A zoomed-in view of OCT B-scans provides the outer retinal profile (red line) from which the OS length is obtained and plotted as a function of eccentricity in a color map (B). Warmer and cooler colors represent longer and shorter OS lengths respectively. Retinal function is assessed by measuring the change in outer segment optical path length (ΔOPL) evoked by a light stimulus (shaded green) – denoted as the ORG amplitude (D). The saturated ORG amplitude is obtained from the time evolution of the functional response, shown in (D) for a near-foveal (gray) and parafoveal (black) retinal location. The shaded areas around the solid lines indicate the standard-deviation of repeat measurements. The variation of the saturated ORG is plotted as a function of eccentricity in a color map (E). Test-retest (left) and Bland-Altman analysis (right) shows high repeatability for both OS length (C) and ORG (F).

### ORG: imaging system, protocol and analysis

ORG was acquired using a custom-built, line-scan spectral domain OCT system with 3° × 5.5° field-of-view, whose details are described previously ^56^. First, after a 1-minute dark adaptation, fifty OCT volumes were recorded at 32 Hz. Single OCT volumes were reconstructed and registered with each other to compensate for small eye movements, rendering an averaged, high signal-to-noise ratio OCT volume (Fig 1A) for further analysis. Next, segmentation of the outer retinal layers – the inner-outer segment (ISOS)-ellipsoid band and the outer segment tips, yielded *en face* 2D images of these layers (Fig 1A) as well as the length of the cone outer segment (Fig 1B). Although EZ width is a well-recognized clinical biomarker of RP, it reflects the preserved portion of the photoreceptor inner segment and therefore does not provide information on outer segment integrity. Sub-clinical changes, such as shortening of the outer segment, can still occur within the EZ area in RP. Therefore, in addition to EZ width (Table 1) derived from clinical OCT based on the presence of the ISOS-ellipsoid band (as determined by three independent graders), this study also used the OS length as a structural measure of disease status. Fig 1B shows the characteristic decline in OS length radiating outward from the foveal center. Note that the measures of cone structure and function, OS length, EZ width and ORG, were all obtained from the same retinal location for each follow-up visit, facilitating spatially localized comparison between them.

The cone ORG, i.e. light-evoked activity in cone OS, was measured following a green stimulus (532 ± 5 nm) of photon density 14.1×10^6^ photons/µm^2^, chosen to provide a similar *and* high activation of L and M-cones. The cone ORG response is defined as the change in the cone OS length evoked by a light stimulus as a function of time, as illustrated in Fig. 1C. It is computed as the change in optical phase (converted to optical path length (ΔOPL)) between the ISOS-EZ and cone outer segment tips (COST). In the typical single-flash stimulus paradigm shown here, the ΔOPL exhibits a rapid, low amplitude reduction immediately after stimulus onset, followed by a sharp increase and a saturated plateau (Fig. 1C). Saturated ORG response is calculated as the average ΔOPL over ∼300 ms during the plateau, as shown in Fig. 1D. In a typical healthy control, the saturated ORG scales linearly with OS length (and eccentricity), whereby longer foveal OS exhibits larger ORG responses and shorter peripheral cones have proportionally reduced ORG^56^, as shown in Fig. 1B&D. The entire ORG procedure was completed in 25 ± 10 mins.

### Test-retest variability and statistical analysis

Figures 1C&F illustrate scatter plots of test-retest variability (left) for OS length and ORG in normal controls, showing minimal changes over two visits, spaced 6-months apart. Each scatter plot includes a linear fit line, which closely approximates the identity line (*y = x*), indicating strong agreement between measurements from the two visits. Figures 1C&F display Bland-Altman plots (right) for OS length and ORG. In these plots, the mean difference (bias) between visits is close to zero for both parameters, indicating no significant systematic bias. Most data points fall within the limits of agreement (±1.96 SD), suggesting minor measurement variability for both OS length and ORG. To further assess the reliability and repeatability, test-retest variability was analyzed by calculating the coefficient of variation (CV) (CV = (SD/Mean) ×100%) for EZ width, OS length, and ORG across repeat measurements. Additionally, Wilcoxon signed-rank tests were used to examine intra-subject variability in reductions of these metrics, providing a means to analyze paired differences for non-normally distributed data. The CVs of 1.5% for EZ width, 3.0% for OS length, and 4.0% for ORG, confirm comparable measurement reliability across these metrics.

## Results

Table 1 lists age, sex, genetic diagnosis, clinical presentations, follow-up duration, and changes in BCVA and EZ width of the RP participants included in the study. Over the course of follow up, there was minimal change in BCVA (Table 1). The EZ width captured on a commercial OCT macular B-scan revealed variable progression across the cohort. Study participants experienced a reduction in EZ width ranging from 0.9% to 15% (mean: 4.5% ± 5.9%) over varying follow-up periods of 12 to 21 months.

The most significant EZ width progression of 15% over 20 months of follow-up was seen in Subject RP01. We detail the complete procedure, including both the clinical and advanced imaging with this subject as an example (Fig. 2). Clinical OCT B-scans and FAF images in Fig. 2 (A-B) illustrate visible changes between baseline and follow-up visits. Within the EZ area, where subtle and early changes might not be distinguishable using clinical biomarkers alone, ORG imaging was applied to an area aligned with the FAF (blue rectangle, white ellipse, Fig 2B & 2C). At baseline, near the border of the healthy island and transition zone (yellow ellipse), a complete loss of OS tip reflection precludes reliable OS length and ORG measurements. The OS length map shows a radial decrease from the fovea to the periphery both at baseline and follow-up.

**Figure 2.**
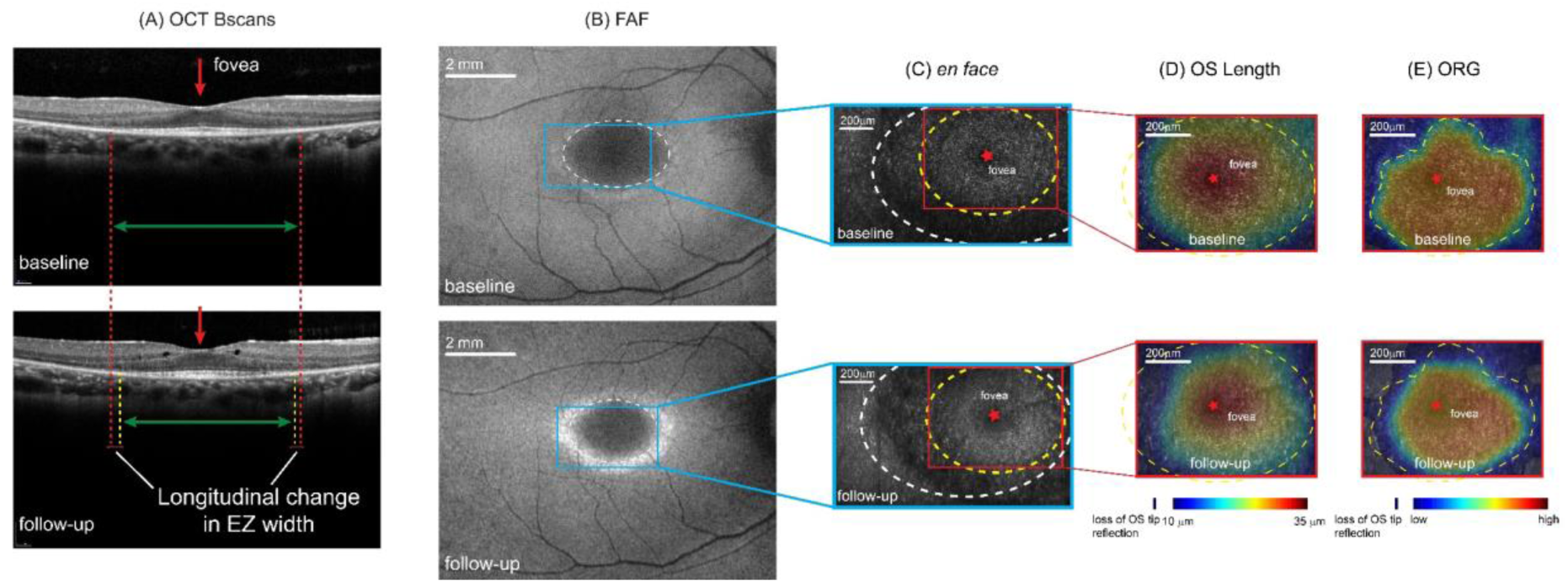
Example longitudinal study analysis pipeline for RP01. (A) Clinical OCT B-scans from baseline (top) and follow-up (bottom) visit. Red arrows indicate the fovea. Red & yellow dashed lines mark the EZ width boundaries at baseline and follow-up respectively. (B) Fundus autofluorescence (FAF) from baseline (top) and follow-up (bottom) visit. The white dashed ellipse outlines the EZ area, while the blue rectangle marks the region selected for further OCT-ORG analysis. (C) E*n face* images for baseline (top) and follow-up (bottom), with the corresponding EZ area from (B) marked as a white dashed ellipse. The yellow ellipse highlights the area chosen for OSL and ORG analysis, based on the preservation of all outer retinal layers. Maps of (D) OS length and (E) ORG amplitude are shown for baseline (top) and follow-up (bottom). An outline is drawn in the OS length and ORG maps, as reference for comparison between baseline and follow-up visits.

However, at follow-up, a reduction in OS length is noted at locations close to the edge of the healthy zone, that is greater in magnitude for the temporal retina. The ORG map follow-up reflects the patient’s general trend of overall reduction in OS length, with the temporal meridian showing the greater progression. In both OS length and ORG, the progression is expectedly most evident at the frontier of degeneration at the border of the healthy island and transition zone. Together, figures 2D-E demonstrate that, even in healthy regions appearing intact on FAF and OCT, advanced ORG imaging not only reveals progressive changes in retinal structure, but also reduced function over time.

Figures 3A-3C display *en face* OCT images segmented at the ISOS-EZ, OS length maps, and ORG amplitude maps for RP03, RP04 and RP05. The red boxes indicate ROIs selected for ORG. The gradual reduction in OS length and ORG versus eccentricity is evident both at baseline and follow-up for all cases, similar to normal controls (Fig 1). In RP03, both OS length and ORG show a significant reduction along the border of the healthy island and transition zone (Fig. 3B, yellow dashed line). The difference in structural and functional readouts are apparent, where the OS length shows a gradual reduction at the edge of the healthy zone, while the ORG shows more drastic decline.

**Figure 3.**
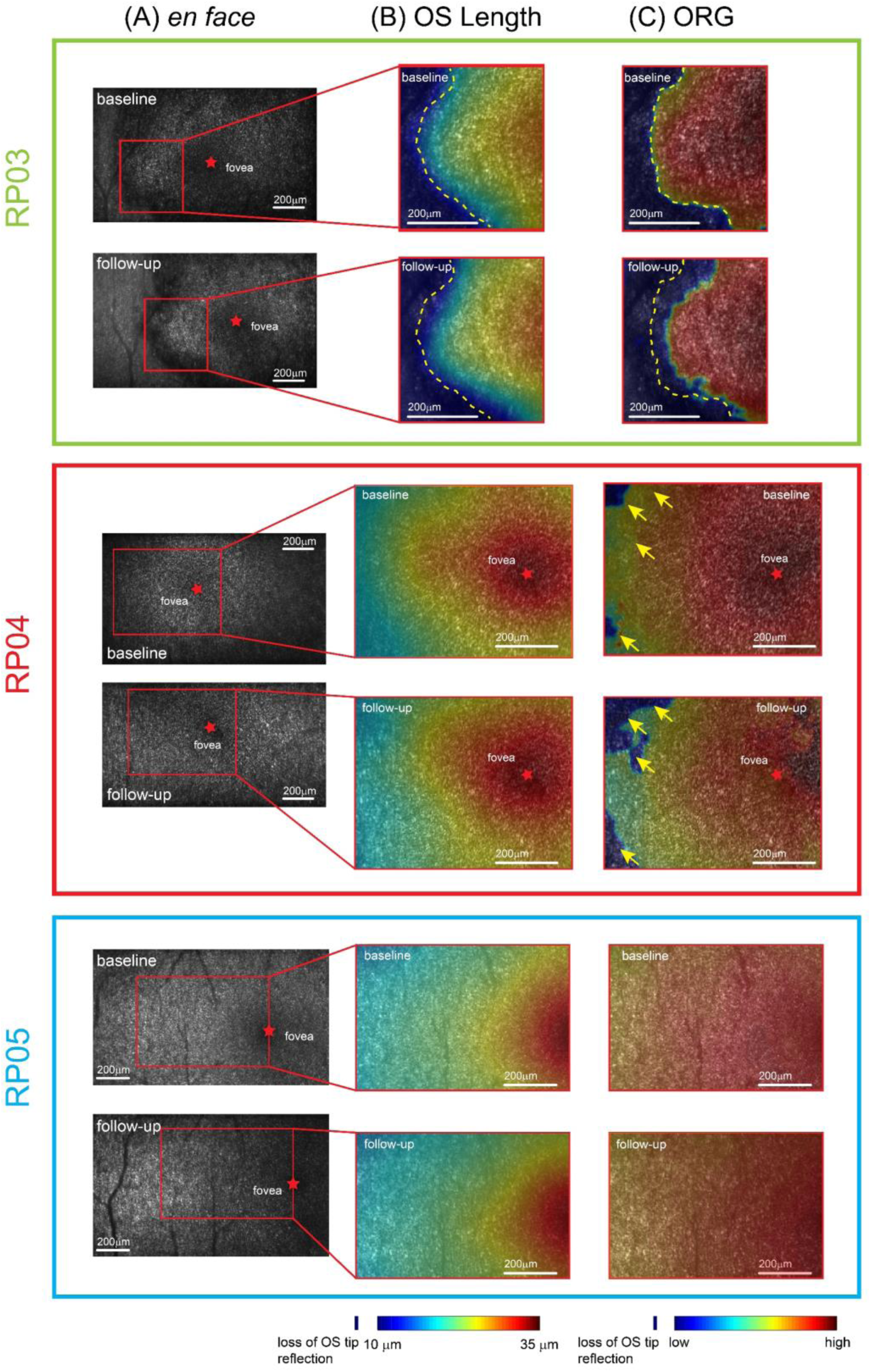
Longitudinal follow-up in RP03, RP04, and RP05, displaying the progression from the initial baseline visit to the most recent follow-up. (A) Baseline and follow-up *en face* images at the ISOS-EZ band and the corresponding ROIs (red) selected for OS length and ORG analysis. Maps of OS length and ORG are shown in (B) and (C) respectively. Their corresponding color bars denote the variation in OS length and ORG response amplitude. Yellow dashed lines and arrows indicate the same location for reference across follow-ups.

In RP04, OS length changes are minor and nearly indistinguishable upon visual inspection of the color maps. Yet ORG reductions are observed, where only minor changes are observed in cone structure as assessed by OS length (yellow arrows, Fig. 3). In the early-stage patient RP05, as defined by minimal symptoms and greater preservation of clinical metrics, both OS length and ORG remained stable over time and no changes were apparent upon visual inspection. Of note, RP03 (age range: 56 - 60 yo) and RP05 (age range: 26 - 30 yo) are both affected by variants in the *RHO* gene (RP03: c.165C>A, p.Asn55Lys; RP05: c.68C>A, p.Pro23His in RP05), but exhibit distinct progression patterns presumably due to their distinct genotypes and disease stage.

The ORG maps reveal a drop in amplitude at the borders of the healthy zone over time, whereas OS length shows comparably lower reductions in the same areas. On the other hand, OS length and ORG remain more stable near the fovea, indicating structural and functional preservation in locations away from the active area of degeneration, i.e. the transition zone. Although the disease progression is minimal near the fovea, the absolute ORG magnitude is indeed decreased in RP compared to normal controls (see Table 2). Previously, the reduction of ORG in the parafovea was shown to correspond with normal microperimetry in RP ^21^. These qualitative observations highlight nuanced structural and functional changes detectable by ORG, even in regions previously considered stable via standard clinical assessment. Next, we performed a quantitative comparison between ORG, OS length and EZ-width as an objective basis for evaluating progression in RP.

**Table 2.**
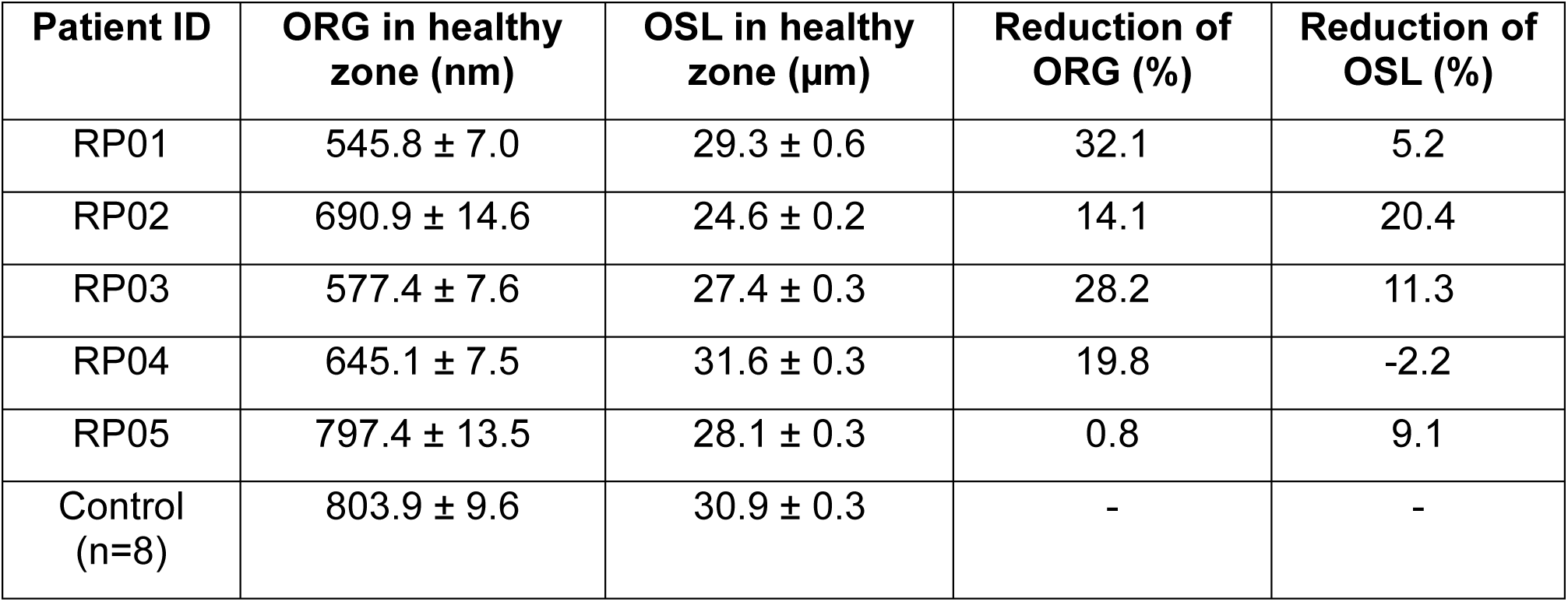
Comparison of ORG and OS length between RP patients and normal controls in the healthy central island.

To derive relevant metrics for disease and potential clinical trials, we divided the ORG into three spatial sections for further assessment: ORG*tot* denoting the overall ORG amplitude across the measurable region, where the measurable region is defined by the presence of both the ISOS-ellipsoid and COST bands; ORG*edge* denoting the ORG response in the border region surrounding the healthy island; and ORG*cent* denoting the ORG response in the central healthy region. This is analogous to the *“hill of vision”* metric ^57,58^ for documenting the rate of progression in established natural history studies such as the *USH2A*-related retinal degeneration or RUSH2A study. ORG*edge* and ORG*cent* were calculated based on the mean ORG of 10 regions of interest, each of size 0.25° × 0.25°, near the border of the healthy island and in the central fovea respectively (marked by white dots in Fig 4A). The ORG*tot* was calculated as the mean ORG response for the whole measurable area (white dashed line in Fig. 4A). The comparison between these spatially distinct metrics is shown in Fig. 4B. ORG*edge* is the most sensitive for assessing progression and demonstrates the largest reduction over time, highlighting the active degradation at the boundary of the transition zone. The corresponding changes in OS length for the same spatial sub-divisions - indicated as OSL*tot*, OSL*edge*, and OSL*cent* - are illustrated in Fig. 4C. The greater reduction in OSL*edge* compared to OSL*tot* and OSL*cent* is evident in RP01, 03 and 04, while this difference is not apparent for the other two cases.

**Figure 4.**
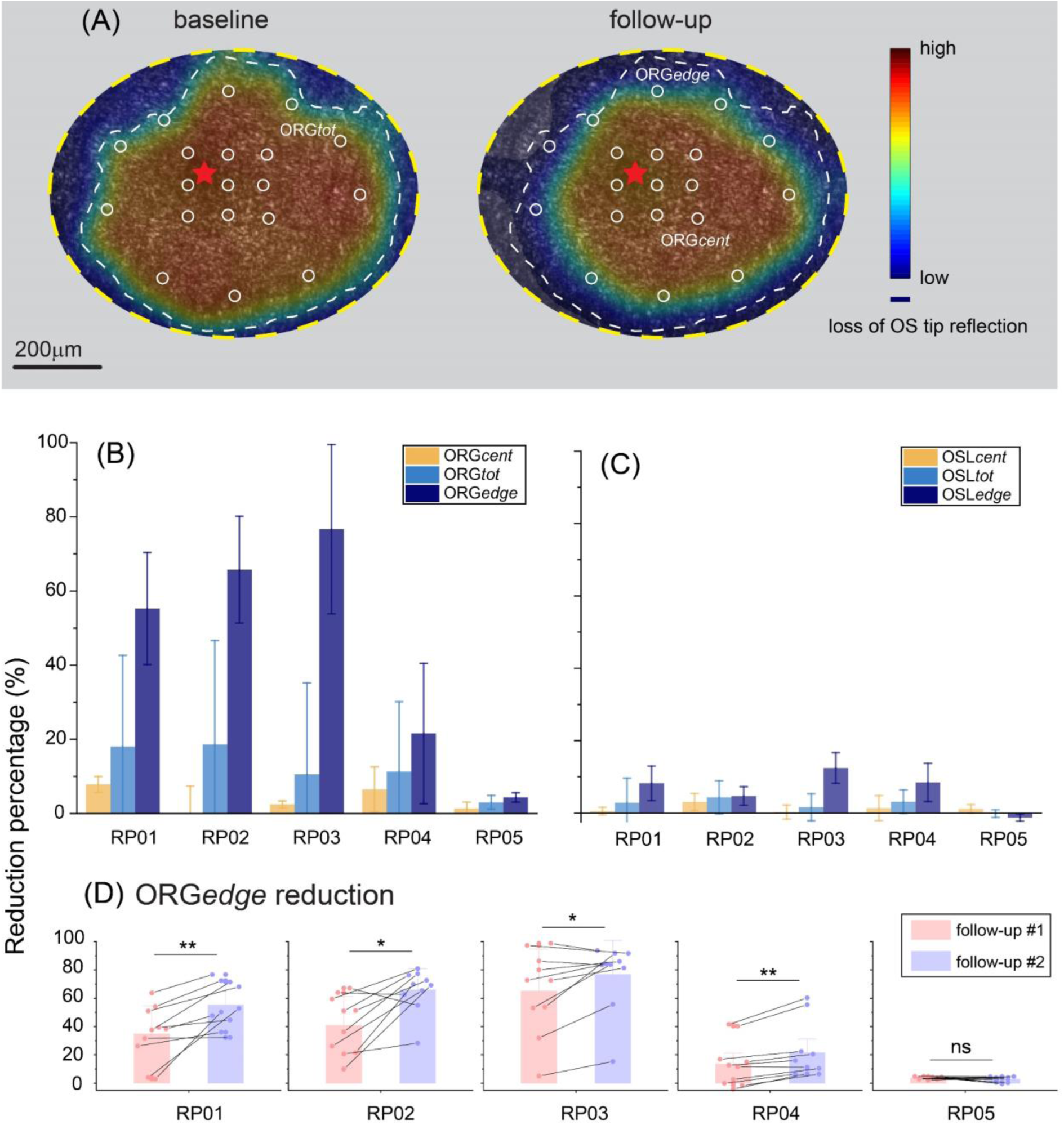
Comparison of biomarkers for detecting RP disease progression. (A) The ORG maps are divided into regions-of-interest (ROIs) (white circles) to allow regional comparison of photoreceptor dysfunction. ORG*edge* and ORG*cent* are derived from ROIs at the border of the healthy island and in the central fovea, respectively, while ORG*tot* is obtained from the entire ORG map. The fovea is indicated by a red star. (B) Comparison of regional metrics derived from ORG, including ORG*tot*, ORG*edge*, and ORG*cent*. (C) Corresponding metrics from OS length, including OSL*tot*, OSL*edge*, and OSL*cent* in (C). (D) Scatter and bar plots showing the percentage reduction in ORG*edge* amplitude over time for the five RP patients, with pink and purple shades representing the first and second follow-up visits, respectively. Connecting lines illustrate ORG changes for the same imaged location across the different follow-ups. Statistical significance values obtained from the Wilcoxon signed-rank test are noted for (D).

With the knowledge that ORG*edge* was the most sensitive in detecting longitudinal changes, we plotted the change of this metric for the two follow-ups (Figure 4D). It was evident that by 9 months after their baseline visit, there was detectable disease progression in RP03 and RP04, as indicated by the reduction in ORG*edge* at the first follow-up – 65.1% and 13.5% respectively. For RP01 and RP02, the first follow up of 16 and 14 months respectively showed an ORG*edge* reduction of 34.9% and 40.7% respectively. There was a notable decrease in ORG*edge* amplitude for all patients at both follow-ups, apart from RP05: f/u #1: mean = 31.5% ± 6.7% (range: 3.6% - 65.1%) and f/u #2: mean = 44.7% ± 5.3% (range: 4.3% - 76.7%). For RP01 – 03, the progression was statistically significant for both follow-ups compared to baseline (p <0.005). For RP04, the progression from baseline to 1^st^ follow-up was not statistically significant (p = 0.084), while the progression from baseline to the 2^nd^ follow-up was significant (p = 0.02). In these four cases, cone function deteriorated significantly in the time-interval between the two follow-up visits, demonstrated by the larger ORG reduction at the second follow-up. The most advanced stage patient within the cohort (RP03) also demonstrated the largest reduction in ORG*edge* (65.1% and 76.7% respectively at 9 and 15-month follow up).

We then compared ORG*edge* to standard clinical metrics of outer retinal structure (Fig. 5). Comparison of the reduction in ORG*edge* against EZ width and OS length for the most recent follow-up visits is shown. Note that OS length in Fig. 5 is obtained from the same locations and regions-of-interest as the ORG*edge*, facilitating a direct comparison between cone structure and function at the edge of the healthy island. Significantly greater reduction in ORG*edge* amplitude was observed compared to OSL and EZ width for RP01, 02, 03 and 04 (p < 0.02, Wilcoxon signed-rank test). For RP05, the small but significant reduction in ORG*edge* was not accompanied by changes in EZ width.

**Figure 5.**
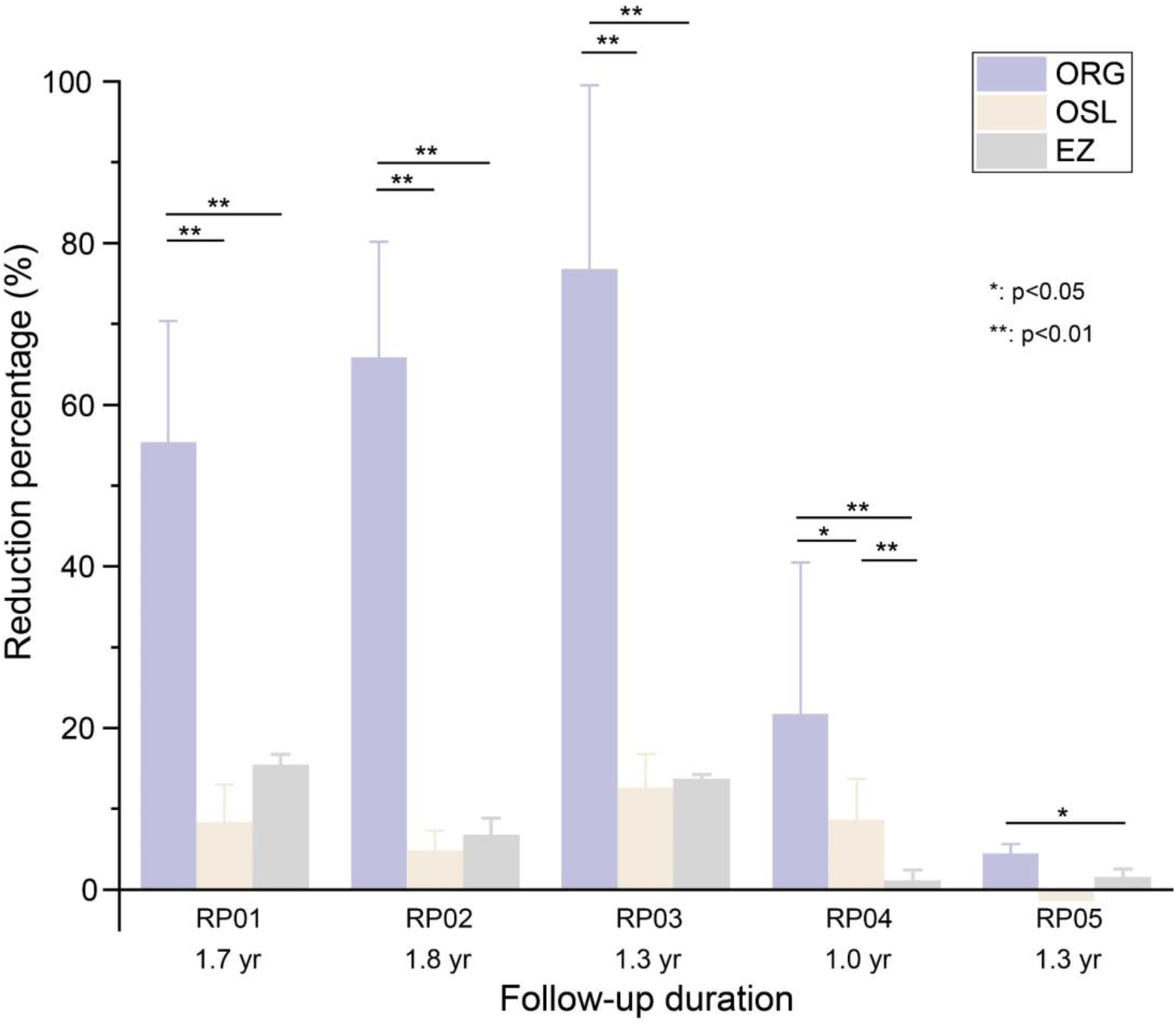
Comparison between ORG*edge*, OS length, and EZ-width for the five RP patients. Statistical significance values obtained from the Wilcoxon signed-rank test are noted in the plot.

Deterioration in cone structure, measured by a reduction in OSL and EZ, was less pronounced; the change in OS length and EZ-width was 5.7% ± 1.2% and 4.5% ± 5.9% respectively. The comparative reductions in EZ width versus OS length were not statistically different except for in RP04. Overall, these findings demonstrate that ORG detection of cone dysfunction precedes the loss of cone structure and provides an early and otherwise sub-clinical biomarker of disease progression.

## Discussion

This is the first study to assess longitudinal disease progression using ORG and highlights its vast potential as a sensitive biomarker to precisely monitor progression of outer retinal disease. While RP was chosen as an example of outer retinal degeneration to assess the sensitivity of ORG to detect progression, the findings could be applicable across a broad range of primary and secondary photoreceptor diseases, including geographic atrophy in age-related macular degeneration (AMD) and other inherited retinal diseases, where the structure and function of the outer retinal complex is compromised ^13,59,60^. The evolution of ORG used in this study, dubbed CoORG or Coarse-scale ORG, does not require an adaptive optics module and sacrifices cellular resolution for increased field-of-view and high-throughput, patient-friendly imaging ^56^. Furthermore, patients with characteristics such as cataracts and excessive eye movements that would preclude adaptive optics imaging are accessible within the CoORG paradigm. Longitudinal study of the same patients and retinal locations over time as demonstrated here offers inherent control over inter- and intra-subject dependent factors that cause variability in the ORG, facilitating effective comparisons. In the future, documenting the normative characteristics of the ORG versus eccentricity ^56^ and age are important considerations for its wide-spread application.

Disease progression measured in ORG was compared to standard clinical assays. EZ width in OCT is one of the key biomarkers that reflects the remnant inner-segment-EZ structure of the photoreceptors. Here, we observed a 5% annual decline in EZ width in our cohort, a finding that closely aligns with the 5-7% reduction rate reported in previous studies ^6,11^. While EZ width is a highly repeatable marker of advanced-stage outer retinal degeneration ^7,55^, it does not convey functional information of the outer retina, nor the viability of the photoreceptor outer segment. The constriction of hyper-AF rings observed in FAF provides a biomarker for progression and is shown to have low test-retest variability ^55^. In RP, the annual horizontal and vertical rates of decline in hyper-AF ring size were reported to be 4.1% and 4.0%, respectively ^55^. In our study, we observed similar rates of annual decline over a 5-year period for RP01, with horizontal and vertical reductions of 4.1% and 3.4%, respectively.

As an add-on to OCT imaging, the ORG seamlessly integrates the assessment of structure and function in the same platform with high sensitivity and spatial localization, enabling the detection of functional changes that are anchored to clinically relevant structural disease phenotypes such as EZ width and OS length. This is of critical importance not only to compare against standard clinical measures of retinal structure but also to obtain granular insight into the spatially heterogeneous pattern of disease progression, such as in sectoral RP or geographic atrophy. As expected from the typical pattern of progression in RP, from the periphery to the fovea, the highly variable border region near the healthy island denoted by ORG*edge* showed the largest reduction over time. In contrast, cone function in the central healthy area, denoted by ORG*cent*, showed minimal change over time. (Figure 4B-C). However, cone function in the central retina appeared to be compromised compared to controls even at the first time point, as shown in Table 2, consistent with cross-sectional reports of ORG in RP ^21,39^. The ORG*tot* shows intermediate reduction in cone function compared to ORG*edge* and ORG*cent* (Figure 4B). Overall, targeted regional analysis as introduced here for ORG, based on previous suggestions for visual sensitivity ^57,58^, enables capturing the spatially localized and heterogenous pattern of disease progression as well as its comparison against more global clinical metrics.

As shown in Fig 5, the reduction in cone function with ORG over time exceeds that observed in EZ width and OS length. EZ width and OSL decreased by 4.5% ± 5.9% and 6.5% ± 1.4%, respectively (Fig. 5), approximately 9.9 and 6.9 times less than the reduction noted in ORG*edge*. The ORG*edge* revealed an average reduction of 44.7% ± 5.3% including the very early-stage subject (RP05). If RP05 were excluded, the mean ORG reduction in the remaining patients would increase to 54.8% ± 3.1%. The comparison against ORG*tot* was more modest, where reduction in overall cone function with ORG exceeds reductions in EZ width and OS length by 2.7x and 1.9x respectively (12.3% for ORG vs. 4.5% for EZ width and 6.5% for OS length). Whereas the EZ width can be treated as global metric of outer retinal health encompassing both the central healthy island and the transition zone, the breakdown of OS length into *edge*, *center* and *total* allows a more regional comparison between structure and function. Comparing Figure 4B versus Figure 4C, it is evident that reduction in cone function over time generally exceeds the reduction in OS length, whether assessed at the edge of the healthy island, or as a whole.

The growing landscape of clinical trials, including the advancements in novel gene-augmentation and gene-agnostic therapies for outer retinal diseases, underscores the need for outcome measures that are safe, sensitive, and efficient. These measures are particularly critical at early time points, when interventions may be most effective, or when the therapeutic effects can be assessed earlier during a clinical trial. The approval of voretigene neparvovec (Luxturna) for patients with *RPE65* mutations causing Leber Congenital Amaurosis (LCA) represented a major advance in human vision restoration using gene therapy. In addition, gene-editing for *CEP290*-associated retinal degeneration has shown promising early results in clinical trials. In the context of geographic atrophy in AMD, pegcetacoplan and avacincaptad pegol drugs have recently received FDA approval for slowing disease progression by targeting the complement pathway ^61–63^. EZ-width and FAF are commonly used as clinical trial outcome measures. Efforts aimed at slowing or reversing disease progression would benefit from a faster and more sensitive biomarker that could ultimately lead to a shorter pathway for therapeutic testing and approval. This study marks a significant step in this direction. The low test-retest variability (∼4%) coupled with the high sensitivity in detecting annual rates of reduction in cone function highlights the potential of ORG to augment longitudinal studies of retinal disease and response to therapeutic interventions. Accordingly, for a treatment to show measurable effect with ORG, it would need to exhibit a minimum of 4% preservation or restoration of cone function, while a treatment effect would be measurable in a significantly shorter timespan compared to the current outcome measures.

### Limitations

The study sample size is small, which may constrain statistical power and generalizability, specifically with respect to drawing genotype-phenotype correlations. In addition, the longitudinal study duration is limited to 21 months. Nevertheless, the ORG measurements showed a high test-retest reliability over repeat measurements, and significant disease progression was observed with ORG, even within the relatively short timeframe and in those with limited progression with standard clinical imaging. Another limitation of our study is the lack of longitudinal microperimetry in the same subjects. The addition of this important biomarker would enable relating the change in visual field to the degradation noted here in ORG and retinal structure. Previous work shows that retinal areas with sub-normal cone function measured with ORG ^21^ may exhibit normal visual sensitivity in RP, while IRD patients with loss of up to ∼62% of cones still exhibit normal visual acuity and sensitivity ^15^. This suggests that subjective tests of visual function may be less sensitive compared to objective tests of cone function such as ORG in tracking progression.

## Conclusions

Overall, this study highlights the ability of ORG to capture early changes in photoreceptor function over time that are otherwise undetectable in standard clinical imaging and often precede structural deterioration. By enabling the detection of functional impairments before structural changes become apparent, ORG is poised to play a pivotal role in clinical practice and therapeutic trials. As treatments continue to evolve, the incorporation of ORG into clinical trials will accelerate the timeline and improve sensitivity for monitoring treatment efficacy, ultimately shortening the assessment period and enhancing patient outcomes.

## Data Availability

All data produced in the present study are available upon reasonable request to the authors.

## Acknowledgements/Disclosure

The following sources of funding are acknowledged: NIH grant U01EY032055, EY029710, Unrestricted grant from the Research to Prevent Blindness, Dawn’s Light Foundation, George and Martina Kren Professorship in Vision Research. Vimal Pandiyan and Ramkumar Sabesan have filed a patent on the line-scan OCT technology used here for optoretinography (PCT/US2020/029984).

## Notes

### Funding Statement

This study was funded by:
NIH grant U01EY032055, EY029710, Unrestricted grant from the Research to Prevent Blindness, Dawn's Light Foundation, George and Martina Kren Professorship in Vision Research.

### Author Declarations

The institutional review board of the University of Washington (IRB#STUDY00002923) gave ethical approval for this work.

### Summary of Updates

The website author affiliations updated. Just to make sure the doi's author affiliations is consistent with the manuscript we submitted previously.

## References

1. Russell S, Bennett J, Wellman JA, et al. Efficacy and safety of voretigene neparvovec (AAV2-hRPE65v2) in patients with RPE65-mediated inherited retinal dystrophy: a randomised, controlled, open-label, phase 3 trial. The Lancet. 2017;390(10097):849–860. doi:10.1016/S0140-6736(17)31868-8

2. Schmetterer L, Scholl H, Garhöfer G, et al. Endpoints for clinical trials in ophthalmology. Progress in Retinal and Eye Research. 2023;97:101160. 10.1016/j.preteyeres.2022.101160

3. Scalabrino ML, Thapa M, Wang T, Sampath AP, Chen J, Field GD. Late gene therapy limits the restoration of retinal function in a mouse model of retinitis pigmentosa. Nature Communications. 2023;14(1):8256.

4. Tao LW, Wu Z, Guymer RH, Luu CD. Ellipsoid zone on optical coherence tomography: a review. Clinical & experimental ophthalmology. 2016;44(5):422–430. doi: 10.1111/ceo.12685

5. Scoles D, Flatter JA, Cooper RF, et al. Assessing photoreceptor structure associated with ellipsoid zone disruptions visualized with optical coherence tomography. Retina. 2016;36(1):91–103. doi:10.1097/IAE.0000000000000618

6. Birch DG, Locke KG, Wen Y, Locke KI, Hoffman DR, Hood DC. Spectral-domain optical coherence tomography measures of outer segment layer progression in patients with X-linked retinitis pigmentosa. JAMA ophthalmology. 2013;131(9):1143–1150. doi:10.1001/jamaophthalmol.2013.4160

7. Hariri AH, Zhang HY, Ho A, et al. Quantification of ellipsoid zone changes in retinitis pigmentosa using en face spectral domain–optical coherence tomography. JAMA ophthalmology. 2016;134(6):628–635. doi:10.1001/jamaophthalmol.2016.0502

8. Birch DG, Locke KG, Felius J, et al. Rates of decline in regions of the visual field defined by frequency-domain optical coherence tomography in patients with RPGR-mediated X-linked retinitis pigmentosa. Ophthalmology. 2015;122(4):833–839. 10.1016/j.ophtha.2014.11.005

9. Delori FC, Dorey CK, Staurenghi G, Arend O, Goger DG, Weiter JJ. In vivo fluorescence of the ocular fundus exhibits retinal pigment epithelium lipofuscin characteristics. Investigative ophthalmology & visual science. 1995;36(3):718–729.

10. Robson AG, Tufail A, Fitzke F, et al. Serial imaging and structure-function correlates of high-density rings of fundus autofluorescence in retinitis pigmentosa. Retina. 2011;31(8):1670–1679. doi:10.1097/IAE.0b013e318206d155

11. Cabral T, Sengillo JD, Duong JK, et al. Retrospective analysis of structural disease progression in retinitis pigmentosa utilizing multimodal imaging. Scientific Reports. 2017;7(1):10347. doi:10.1038/s41598-017-10473-0

12. Pole C, Ameri H. Fundus autofluorescence and clinical applications. Journal of Ophthalmic & Vision Research. 2021;16(3):432. doi:10.18502/jovr.v16i3.9439

13. Cleland SC, Konda SM, Danis RP, et al. Quantification of geographic atrophy using spectral domain OCT in age-related macular degeneration. Ophthalmology Retina. 2021;5(1):41–48. 10.1016/j.oret.2020.07.006

14. Vámos R, Tátrai E, Németh J, Holder GE, DeBuc DC, Somfai GM. The structure and function of the macula in patients with advanced retinitis pigmentosa. Investigative ophthalmology & visual science. 2011;52(11):8425–8432. 10.1167/iovs.11-7780

15. Ratnam K, Carroll J, Porco TC, Duncan JL, Roorda A. Relationship between foveal cone structure and clinical measures of visual function in patients with inherited retinal degenerations. Investigative ophthalmology & visual science. 2013;54(8):5836–5847. 10.1167/iovs.13-12557

16. Battaglia Parodi M, La Spina C, Triolo G, et al. Correlation of SD-OCT findings and visual function in patients with retinitis pigmentosa. Graefe’s Archive for Clinical and Experimental Ophthalmology. 2016;254:1275–1279. 10.1007/s00417-015-3185-x

17. Grover S, Fishman GA, Brown Jr J. Patterns of visual field progression in patients with retinitis pigmentosa. Ophthalmology. 1998;105(6):1069–1075. 10.1016/S0161-6420(98)96009-2

18. Holder GE. Pattern electroretinography (PERG) and an integrated approach to visual pathway diagnosis. Progress in retinal and eye research. 2001;20(4):531–561. 10.1016/S1350-9462(00)00030-6

19. Birch DG, Anderson JL, Fish GE. Yearly rates of rod and cone functional loss in retinitis pigmentosa and cone-rod dystrophy. Ophthalmology. 1999;106(2):258–268. 10.1016/S0161-6420(99)90064-7

20. Lee JY, Care RA, Kastner DB, Della Santina L, Dunn FA. Inhibition, but not excitation, recovers from partial cone loss with greater spatiotemporal integration, synapse density, and frequency. Cell reports. 2022;38(5)doi:10.1016/j.celrep.2022.110317

21. Wendel BJ, Pandiyan VP, Liu T, et al. Multimodal High-Resolution Imaging in Retinitis Pigmentosa: A Comparison Between Optoretinography, Cone Density, and Visual Sensitivity. Invest Ophthalmol Vis Sci. Aug 1 2024;65(10):45. doi:10.1167/iovs.65.10.45

22. Gerth C, Wright T, Héon E, Westall CA. Assessment of central retinal function in patients with advanced retinitis pigmentosa. Investigative ophthalmology & visual science. 2007;48(3):1312–1318. 10.1167/iovs.06-0630

23. Pierce EA, Aleman TS, Jayasundera KT, et al. Gene editing for CEP290-associated retinal degeneration. N Engl J Med. 2024;390:1972–1984. doi:10.1056/NEJMoa2309915

24. Lorenz B, Künzel SH, Preising MN, et al. Single Center Experience with Voretigene Neparvovec Gene Augmentation Therapy in RPE65 Mutation–Associated Inherited Retinal Degeneration in a Clinical Setting. Ophthalmology. 2024;131(2):161–178. 10.1016/j.ophtha.2023.09.006

25. Van Gelder RN, Chiang MF, Dyer MA, et al. Regenerative and restorative medicine for eye disease. Nature medicine. 2022;28(6):1149–1156. 10.1038/s41591-022-01862-8

26. Roska B, Sahel J-A. Restoring vision. Nature. 2018;557(7705):359-367. 10.1038/s41586-018-0076-4

27. Levin LA, Chiang MF, Dyer MA, et al. Translational roadmap for regenerative therapies of eye disease. Med. 2023;4(9):583–590. 10.1016/j.medj.2023.06.005

28. Rosin B, Banin E, Sahel J-A. Current status of clinical trials design and outcomes in retinal gene therapy. Cold Spring Harbor Perspectives in Medicine. 2024;14(7):a041301. doi:10.1101/cshperspect.a041301

29. Hillmann D, Spahr H, Pfäffle C, Sudkamp H, Franke G, Hüttmann G. In vivo optical imaging of physiological responses to photostimulation in human photoreceptors. Proceedings of the National Academy of Sciences. 2016;113(46):13138–13143. 10.1073/pnas.1606428113

30. Cooper RF, Tuten WS, Dubra A, Brainard DH, Morgan JI. Non-invasive assessment of human cone photoreceptor function. Biomedical optics express. 2017;8(11):5098–5112. 10.1364/BOE.8.005098

31. Zhang P, Zawadzki RJ, Goswami M, et al. In vivo optophysiology reveals that G-protein activation triggers osmotic swelling and increased light scattering of rod photoreceptors. Proceedings of the National Academy of Sciences. 2017;114(14):E2937–E2946. 10.1073/pnas.1620572114

32. Zhang F, Kurokawa K, Lassoued A, Crowell JA, Miller DT. Cone photoreceptor classification in the living human eye from photostimulation-induced phase dynamics. Proceedings of the National Academy of Sciences. 2019;116(16):7951–7956. 10.1073/pnas.1816360116

33. Ma G, Son T, Kim T-H, Yao X. Functional optoretinography: concurrent OCT monitoring of intrinsic signal amplitude and phase dynamics in human photoreceptors. Biomedical Optics Express. 2021;12(5):2661–2669. 10.1364/BOE.423733

34. Pandiyan VP, Maloney-Bertelli A, Kuchenbecker JA, et al. The optoretinogram reveals the primary steps of phototransduction in the living human eye. Science advances. 2020;6(37):eabc1124. doi:10.1126/sciadv.abc1124

35. Azimipour M, Valente D, Vienola KV, Werner JS, Zawadzki RJ, Jonnal RS. Optoretinogram: optical measurement of human cone and rod photoreceptor responses to light. Optics letters. 2020;45(17):4658–4661. 10.1364/OL.398868

36. Tomczewski S, Węgrzyn P, Borycki D, Auksorius E, Wojtkowski M, Curatolo A. Light-adapted flicker optoretinograms captured with a spatio-temporal optical coherence-tomography (STOC-T) system. Biomedical Optics Express. 2022;13(4):2186–2201. 10.1364/BOE.444567

37. Wongchaisuwat N, Amato A, Yang P, et al. Optical Coherence Tomography Split-Spectrum Amplitude-Decorrelation Optoretinography Detects Early Central Cone Photoreceptor Dysfunction in Retinal Dystrophies. Translational Vision Science & Technology. 2024;13(10):5–5. 10.1167/tvst.13.10.5

38. Pandiyan VP, Nguyen PT, Pugh Jr EN, Sabesan R. Human cone elongation responses can be explained by photoactivated cone opsin and membrane swelling and osmotic response to phosphate produced by RGS9-catalyzed GTPase. Proceedings of the National Academy of Sciences. 2022;119(39):e2202485119. 10.1073/pnas.2202485119

39. Lassoued A, Zhang F, Kurokawa K, et al. Cone photoreceptor dysfunction in retinitis pigmentosa revealed by optoretinography. Proceedings of the National Academy of Sciences. 2021;118(47):e2107444118. 10.1073/pnas.2107444118

40. Gaffney M, Connor TB, Cooper RF. Intensity-based optoretinography reveals sub-clinical deficits in cone function in retinitis pigmentosa. Frontiers in Ophthalmology. 2024;410.3389/fopht.2024.1373549

41. Busskamp V, Picaud S, Sahel J-A, Roska B. Optogenetic therapy for retinitis pigmentosa. Gene therapy. 2012;19(2):169–175. 10.1038/gt.2011.155

42. Maguire AM, Bennett J, Aleman EM, Leroy BP, Aleman TS. Clinical perspective: treating RPE65-associated retinal dystrophy. Molecular Therapy. 2021;29(2):442–463. doi:10.1016/j.ymthe.2020.11.029

43. Kwak JJ, Kim HR, Byeon SH. Short-term outcomes of the first in vivo gene therapy for RPE65-mediated retinitis pigmentosa. Yonsei Medical Journal. 2022;63(7):701. 10.3349/ymj.2022.63.7.701

44. Florido A, Vingolo EM, Limoli PG, Cotento L. Mesenchymal stem cells for treatment of retinitis pigmentosa: short review. HSOA JOURNAL OF STEM CELLS RESEARCH, DEVELOPMENT & THERAPY. 2021;7(1)doi:10.24966/SRDT-2060/100066

45. Lam BL, Pennesi ME, Kay CN, et al. Assessment of Visual Function with Cotoretigene Toliparvovec in X-Linked Retinitis Pigmentosa in the Randomized XIRIUS Phase 2/3 Study. Ophthalmology. 2024;10.1016/j.ophtha.2024.02.023

46. Cai CX, Locke KG, Ramachandran R, Birch DG, Hood DC. A comparison of progressive loss of the ellipsoid zone (EZ) band in autosomal dominant and x-linked retinitis pigmentosa. Investigative ophthalmology & visual science. 2014;55(11):7417–7422. 10.1167/iovs.14-15013

47. Tee JJ, Yang Y, Kalitzeos A, Webster A, Bainbridge J, Michaelides M. Natural history study of retinal structure, progression, and symmetry using ellipzoid zone metrics in RPGR-associated retinopathy. American Journal of Ophthalmology. 2019;198:111–123. 10.1016/j.ajo.2018.10.003

48. Micevych PS, Wong J, Zhou H, et al. Cone structure and function in RPGR-and USH2A-associated retinal degeneration. American journal of ophthalmology. 2023;250:1–11. 10.1016/j.ajo.2023.01.006

49. Hood DC, Lin CE, Lazow MA, Locke KG, Zhang X, Birch DG. Thickness of receptor and post-receptor retinal layers in patients with retinitis pigmentosa measured with frequency-domain optical coherence tomography. Investigative ophthalmology & visual science. 2009;50(5):2328–2336. 10.1167/iovs.08-2936

50. Sayo A, Ueno S, Kominami T, et al. Significant relationship of visual field sensitivity in central 10 to thickness of retinal layers in retinitis pigmentosa. Investigative Ophthalmology & Visual Science. 2018;59(8):3469–3475. 10.1167/iovs.18-24635

51. Aizawa S, Mitamura Y, Baba T, Hagiwara A, Ogata K, Yamamoto S. Correlation between visual function and photoreceptor inner/outer segment junction in patients with retinitis pigmentosa. Eye. 2009;23(2):304–308. 10.1038/sj.eye.6703076

52. Cideciyan AV, Jacobson SG. Negative electroretinograms in retinitis pigmentosa. Investigative ophthalmology & visual science. 1993;34(12):3253–3263.

53. Huang W-C, Liu P-K, Wang N-K. Electroretinogram (ERG) to evaluate the retina in cases of retinitis pigmentosa (RP). Retinitis Pigmentosa. Springer; 2022:111–122.

54. Chan H, Brown B. Investigation of retinitis pigmentosa using the multifocal electroretinogram. Ophthalmic and Physiological Optics. 1998;18(4):335–350. 10.1046/j.1475-1313.1998.00374.x

55. Sujirakul T, Lin MK, Duong J, Wei Y, Lopez-Pintado S, Tsang SH. Multimodal imaging of central retinal disease progression in a 2-year mean follow-up of retinitis pigmentosa. American journal of ophthalmology. 2015;160(4):786–798. e4. 10.1016/j.ajo.2015.06.032

56. Jiang X, Liu T, Pandiyan VP, Slezak E, Sabesan R. Coarse-scale optoretinography (CoORG) with extended field-of-view for normative characterization. Biomedical Optics Express. 2022;13(11):5989–6002. 10.1364/BOE.473475

57. Smith TB, Parker M, Steinkamp PN, et al. Structure-function modeling of optical coherence tomography and standard automated perimetry in the retina of patients with autosomal dominant retinitis pigmentosa. PLoS One. 2016;11(2):e0148022. 10.1371/journal.pone.0148022

58. Duncan JL, Liang W, Maguire MG, et al. Baseline visual field findings in the RUSH2A study: associated factors and correlation with other measures of disease severity. American journal of ophthalmology. 2020;219:87–100. 10.1016/j.ajo.2020.05.024

59. Mitchell P, Liew G, Gopinath B, Wong TY. Age-related macular degeneration. The Lancet. 2018;392(10153):1147–1159. doi:10.1016/S0140-6736(18)31550-2

60. Aboshiha J, Dubis AM, Carroll J, Hardcastle AJ, Michaelides M. The cone dysfunction syndromes. British Journal of Ophthalmology. 2016;100(1):115–121. 10.1136/bjophthalmol-2014-306505

61. Chew EY. Complement inhibitors for the treatment of geographic atrophy. The Lancet. 2023;402(10411):1396–1398. doi:10.1016/S0140-6736(23)01844-5

62. Khanani AM, Patel SS, Staurenghi G, et al. Efficacy and safety of avacincaptad pegol in patients with geographic atrophy (GATHER2): 12-month results from a randomised, double-masked, phase 3 trial. The Lancet. 2023;402(10411):1449–1458. 10.1016/S0140-6736(23)01583-0

63. Liao DS, Grossi FV, El Mehdi D, et al. Complement C3 inhibitor pegcetacoplan for geographic atrophy secondary to age-related macular degeneration: a randomized phase 2 trial. Ophthalmology. 2020;127(2):186–195. 10.1016/j.ophtha.2019.07.011

